# Polygenic burden of short tandem repeat expansions promote risk for Alzheimer’s disease

**DOI:** 10.1101/2023.11.16.23298623

**Authors:** Michael H. Guo, Wan-Ping Lee, Badri Vardarajan, Gerard D. Schellenberg, Jennifer Phillips-Cremins

## Abstract

Studies of the genetics of Alzheimer’s disease (AD) have largely focused on single nucleotide variants and short insertions/deletions. However, most of the disease heritability has yet to be uncovered, suggesting that there is substantial genetic risk conferred by other forms of genetic variation. There are over one million short tandem repeats (STRs) in the genome, and their link to AD risk has not been assessed. As pathogenic expansions of STR cause over 30 neurologic diseases, it is important to ascertain whether STRs may also be implicated in AD risk. Here, we genotyped 321,742 polymorphic STR tracts genome-wide using PCR-free whole genome sequencing data from 2,981 individuals (1,489 AD case and 1,492 control individuals). We implemented an approach to identify STR expansions as STRs with tract lengths that are outliers from the population. We then tested for differences in aggregate burden of expansions in case versus control individuals. AD patients had a 1.19-fold increase of STR expansions compared to healthy elderly controls (p=8.27×10^−3^, two-sided Mann Whitney test). Individuals carrying > 30 STR expansions had 3.62-fold higher odds of having AD and had more severe AD neuropathology. AD STR expansions were highly enriched within active promoters in post-mortem hippocampal brain tissues and particularly within SINE-VNTR-Alu (SVA) retrotransposons. Together, these results demonstrate that expanded STRs within active promoter regions of the genome promote risk of AD.

## Introduction

Alzheimer’s disease (AD) is the most common neurodegenerative disorder in the United States and has a growing prevalence in our aging population, yet there is a lack of effective treatments^1^. Delineating the genetic basis of AD is crucial to uncovering the underlying genes and molecular mechanisms and can spur development of more targeted therapies. Despite studies in hundreds of thousands of individuals, most of the genetic risk for AD has yet to be identified^2^. One source of genetic variation that has not been explored in AD is the approximately one million short tandem repeats (STRs) tracts in the human genome, which are classically defined as DNA sequences composed of repeated units of 2-6 bp motifs. STR tract length (i.e., number of repeat units) is highly polymorphic in the population and linked to widespread gene expression changes, thus representing a potential source of functional genetic variation^3–8^. Moreover, pathogenic expansions in the tract lengths of STRs cause over 30 monogenic neurological disorders such as Huntington’s disease^9,10^. Thus, it is important to assess whether STRs may also contribute to the risk of AD.

Pathogenic expansions of specific STR tracts cause >30 monogenic disorders such as fragile X syndrome, myotonic dystrophy, and Huntington’s disease^9,10^. A single pathogenic STR expansion confers the majority of the genetic risk for these disorders. However, the role of STRs in promoting genetic risk for polygenic disorders such as AD is not well understood. Our ability to understand the relationship between STRs and polygenic disorders has been limited by the need for genome-wide measures of STR lengths in large sample sizes. Recent studies in large cohorts have revealed that patients with autism spectrum disorder and schizophrenia carry a higher burden of germline STR expansions^11–13^. These studies suggest that, in contrast to our traditional view of a single STR conferring disease risk for monogenic disorders, many STRs distributed throughout the genome in aggregate can contribute to neuropsychiatric disease risk in a polygenic fashion.

The majority of STRs in the genome are not within genes and the role of these intergenic STRs with disease is less well-understood. While known disease-associated STR tracts are within gene bodies (exons, introns, or untranslated regions)^9^, pathogenic-length expansions of disease-associated STRs has been linked to alterations beyond protein-coding functions, including severe disruption to histone modifications, DNA methylation, and genome folding^14^. Polymorphic changes in intergenic STRs are also known to correlate with expression of nearby genes often in a tissue-specific manner^5,15,16^. Moreover, STRs correlated with gene expression are enriched at transcriptional start sites and colocalize with putative enhancers^5^. Of note, prior studies have also found widespread alterations in the epigenome profiled post-mortem brain tissue from patients with AD^17–19^. Whether the lengths of intergenic STRs are altered in AD and interplay with epigenetic changes has yet to be explored.

Measuring the lengths of STRs is challenging, especially when interrogating across the entire genome in many individuals. The repetitive nature of STRs makes sequencing and downstream processing prone to errors, and their tract lengths often exceed the lengths of traditional short-read next-generation sequencing reads. However, computational tools have been developed to overcome these challenges to infer the lengths of STRs from short-read sequencing data^20–23^, which have enabled genome-wide genotyping of STRs in cohorts of individuals^11–13^. Consequently, they provide a valuable opportunity to test the association of numerous STRs with disease risk across large cohorts of patients and uncover novel links between STRs and disease risk.

In this work, we sought to understand whether STRs may promote genetic risk of AD. We applied cutting-edge computational tools ExpansionHunter^22^ and gangSTR^20^ to genotype STRs genome-wide using PCR-free WGS data from 2981 individuals with and without AD from the Alzheimer’s Disease Sequencing Project (ADSP). We implemented a rigorous approach to identify STR expansions based on having extreme STR tract lengths. Across the genome, we identified 9641 unique STR tracts that had STR expansions in at least one individual. Strikingly, individuals carrying a high burden of expanded STRs had 3.62 higher odds of having AD and had worse AD neuropathology, thus representing one of the strongest genetic effects on AD risk. STR expansions seen in individuals with AD are enriched in promoter regions active in post-mortem hippocampal tissue. These results suggest a model whereby the cumulative effect of multiple STR expansions across the genome promotes genetic risk of AD and generates important insights into the genetic architecture of AD.

## Results

### Genome-wide profiling of STR tracts in an AD cohort

In this study, we utilized a cohort of 2,393 samples (1,213 AD cases and 1,180 controls) from the National Institute of Aging Alzheimer’s Disease Centers (ADC) cohort of the ADSP WGS dataset. These samples have been sequenced on peripheral blood-derived genomic DNA using a PCR-free Illumina whole genome sequencing (WGS) strategy to >30X coverage using 150 bp paired end sequencing reads. We restricted our analyses to individuals of European ancestry (based on both self-reported non-Hispanic White ethnicity and based on genetic ancestry as determined by principal component analysis [PCA] coordinates) (**Supplementary Fig. 1a**). There were no apparent differences between AD cases and controls based on PCA coordinates (**Supplementary Fig. 1b**), sequencing cohort (**Supplementary Fig. 1c**) or sequencing center (**Supplementary Fig. 1d**). To limit potential technical artifacts that could lead to spurious results, we restricted our analysis to samples sequenced to between 30X and 50X coverage genome-wide. There was no statistically significant difference in sequencing coverage between case and control samples (p=0.055, two-tailed Student’s t-test) (**Supplementary Fig. 1e**). Together, these samples represent a deeply phenotyped case/control cohort that has been carefully filtered to mitigate the potential for technical artifacts and confounders.

To understand the role of STRs in AD, we sought to perform an unbiased genome-wide assessment of the relationship between STRs and AD risk (**Fig. 1a**). There are approximately ∼1 million STR tracts in the genome, which is computationally infeasible to apply across thousands of samples using existing methods. To limit the search space for our analyses and reduce computational costs, we focused on STRs that are polymorphic (i.e., vary in length). To identify polymorphic STRs, we first applied the gangSTR algorithm on a catalog of 895,826 STRs across the genome to a subset of 495 individuals of European ancestry from the ADSP (n=246 AD cases and 249 controls). We identified 237,197 STRs that were polymorphic in these 495 samples. We merged this set of 237,197 STRs with 174,262 STRs previously identified to be polymorphic^22^ to result in a union set of 321,742 unique polymorphic STRs (**Supplementary Table 1)**. We used this panel of 321,742 STRs throughout this study to provide genome-wide assessment of polymorphic STRs.

**Figure 1:**
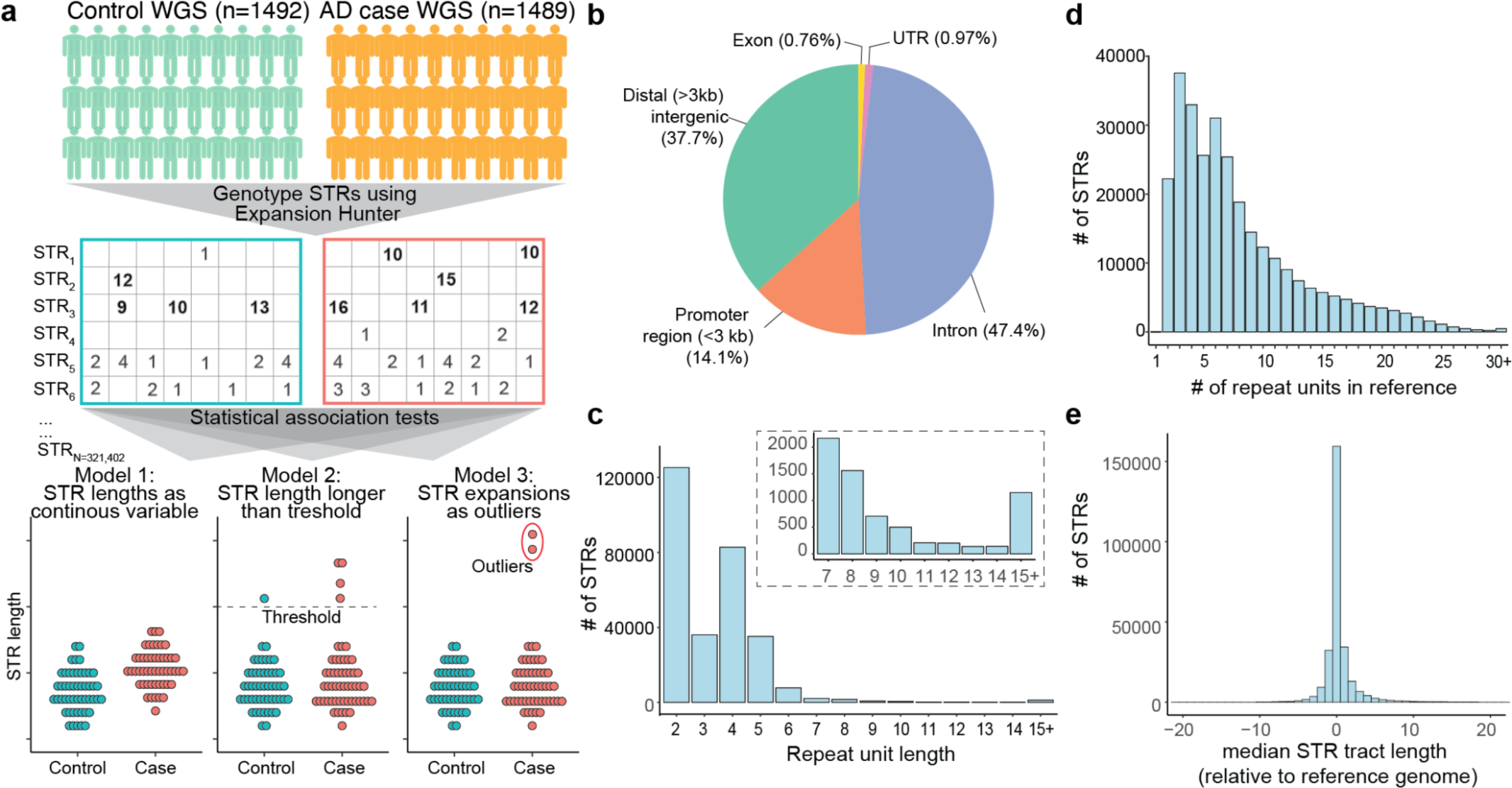
Generation of a panel of polymorphic STRs. **a,** Schematic of study design. Middle section shows six example STRs (STR_1-6_), with rows representing STRs, columns representing individuals, and numbers representing non-reference STR tract lengths. Long expansions are bolded. STR_1_ is an example of a rare STR expansion seen only in individuals with AD. At the bottom are three models for testing for associations with AD case/control status. In model 1 (left), we treat STR tract lengths as a continuous variable. In model 2 (middle), STR expansions are defined as those longer than a given STR tract length threshold, and we compare the number of individuals with a given STR expansion between AD cases and controls. In model 3 (right), we identify rare, long STR expansions. We compare the number of such STR expansions per individual across the genome between cases and controls. **b,** Genomic distribution of STR panel. **c,** Histogram of repeat unit lengths (number of base pairs [bp] in STR motif) for panel. Inset shows the subset of STRs with repeat units 7 bp or longer. **d,** Histogram of STR tract lengths (number of repeat units) in the GRCh38 reference genome for each STR in the panel. **e,** Histogram of median STR tract length relative to the GRCh38 reference genome as genotyped by ExpansionHunter. Negative values indicate a shorter median STR tract length relative to reference genome and positive values indicate longer median tract lengths relative to reference genome.

The STRs in the polymorphic STR set were largely not within protein-coding regions of the genome: 47.4% of STRs were within introns, 36.7% in distal noncoding regions (defined as being outside of gene bodies and at least 3 kb from the nearest transcriptional start site [TSS]), and 14.1% in promoter regions (defined as ≤3 kb upstream from a TSS) (**Fig. 1b**). Only 0.76% of STRs were in coding exons. The high number of polymorphic STRs in gene promoters is consistent with prior studies showing that STRs are enriched in gene promoters and have important gene regulatory roles^24,25^. The majority (97.7%) of STRs had a repeat unit size between 2-6 bp, which is the classic definition of an STR (**Fig. 1c**). Only 0.39% of STRs had a repeat unit size larger than 15 bp. In the GRCh38 reference genome, STRs in the panel generally had short tract lengths, with a median of 6 repeat units per tract and 99.8% of STR tract lengths ≤ 30 repeat units (**Fig. 1d**).

We next used the ExpansionHunter and gangSTR algorithms to genotype our panel of 321,742 polymorphic STRs. We did find overall high concordance between ExpansionHunter and gangSTR genotypes. For 61.2% of STRs, gangSTR and ExpansionHunter genotypes were at least 90% concordant across individuals (**Supplementary Fig. 2a**). The tract lengths were strongly correlated across STRs between gangSTR and ExpansionHunter (Pearson r^2^ 0.83) **(Supplementary Fig. 2b).** We do note that tract lengths in ExpansionHunter tended to be longer than gangSTR, which is consistent with other work showing that ExpansionHunter tends to overestimate STR tract lengths while gangSTR underestimates them^26^. Nonetheless, we found that the median STR tract length measured by ExpansionHunter were largely concordant with the tract lengths in the GRCh38 reference genome (**Fig. 1e**). For the remainder of the paper, we focus on we performed our primary cohort analyses using ExpansionHunter as this algorithm has been shown to be more sensitive for detecting longer STR tract lengths^26^.

### Testing for single STR associations with AD risk

We first tested the lengths of each STR for association with AD risk, treating STR lengths as a continuous variable (**Fig. 1a**, model 1). We applied a logistic regression model to test the association of the tract length of each STR with AD risk, accounting for sample covariates (age, sex, and the first 3 PCs) and technical covariates (sample sequencing coverage, local sequencing coverage, sequencing center). Throughout, we applied a dominant model, such that only the longer of the two alleles for a given STR in an individual was considered. As the tract lengths of many STRs are not normally distributed, we performed a rank-based inverse normal transformation of STR tract lengths prior to association testing. We filtered out STRs within segmental duplicated regions of the genome from our results as these regions resulted in a high number of artifactual genotype calls (see Methods). This resulted in a final set of 293,785 genome-wide STRs that we report. Our statistical association test provided well-calibrated results (λ genomic control=0.997), suggesting minimal evidence of systemic technical artifacts or population stratification (**Fig. 2a; Supplementary Table 2**). We identified one STR associated with a clinical diagnosis of AD at a Bonferroni-corrected p-value threshold of 1.70 x 10^−7^. This STR was a TTTA repeat at chr19:44921097-44921125 (p-value 4.29 x 10^−10^, logistic regression test) and is located approximately 11.7 kb downstream from the *APOE* gene, which has the strongest known genetic association with AD risk^27^.

**Figure 2:**
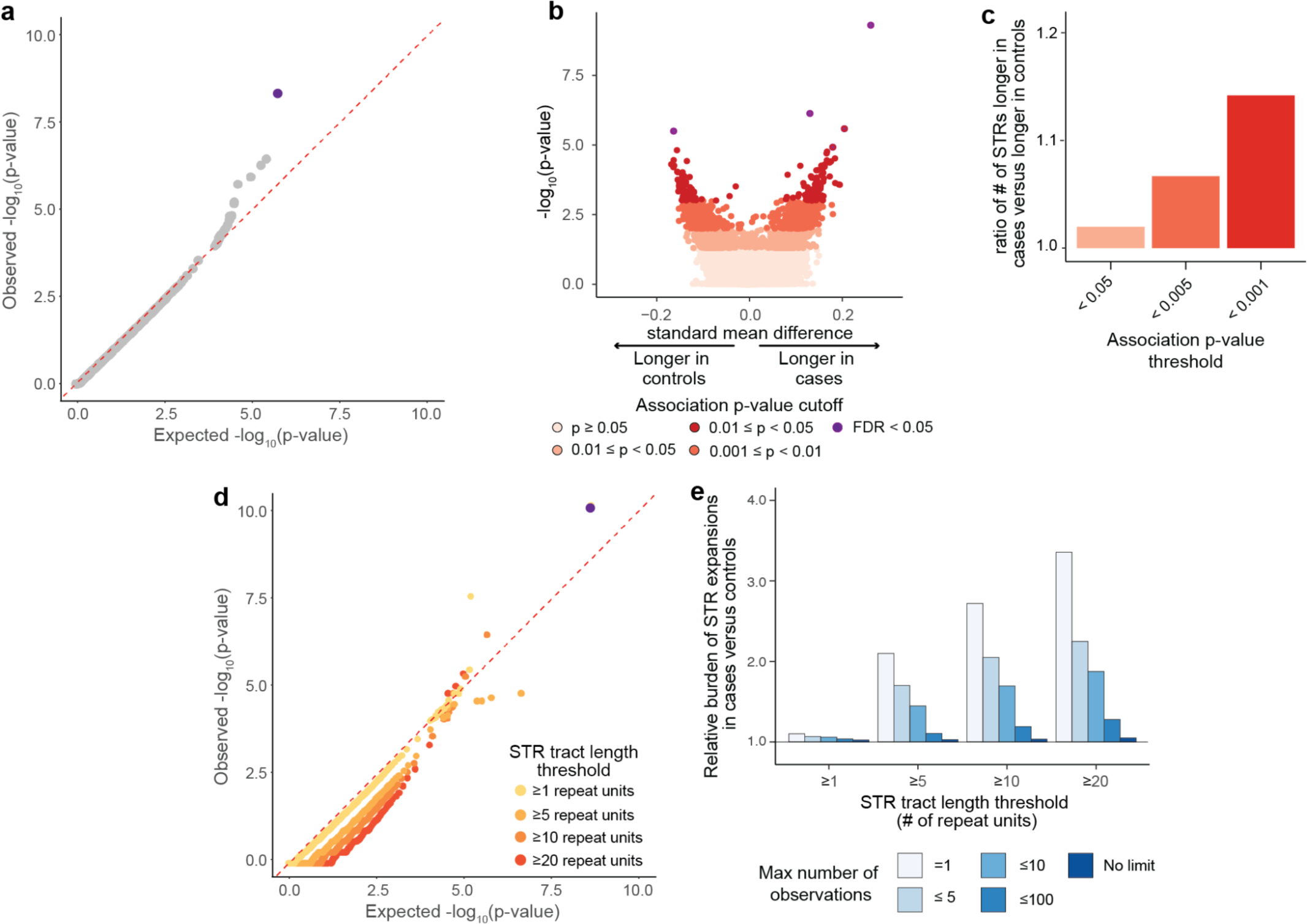
Statistical testing for single STR associations with AD risk. **a,** Quantile-quantile plot of single STR association statistics. X-axis shows the expected distribution of −log_10_(p-values) under a uniform p-value distribution. Y-axis shows observed −log_10_(p-values). Each point represents a separate STR. **b,** Volcano plot for STR tract lengths under a single STR association test. Each point represents a single STR, with colors reflecting statistical significance of association. X-axis reflects standard mean difference, which is the difference in mean STR lengths between cases and controls divided by the standard deviation of STR lengths across the whole cohort. Positive values reflect longer mean STR tract lengths in cases as compared with controls. **c,** Ratio of number of STRs with tract lengths that are longer in cases relative to the number of STRs with tract lengths longer in controls at different statistical significance thresholds. **d,** Quantile-quantile plot of p-values hypergeometric test comparing number of STR expansions in cases versus controls for each STR. X-axis shows the expected distribution of −log_10_(p-values) under a uniform p-value distribution. Y-axis shows observed −log_10_(p-values). Each point represents a separate STR, and points are colored by the STR tract length threshold. **e,** Relative burden of STR expansions in case versus control individuals at different STR tract length and frequency thresholds. Values above 1.0 reflect higher burden of expansions observed in cases than controls.

We performed several validation steps for our association analysis. We first repeated the association analysis using two alternative statistical models. To ensure that associations do not result from the inverse normal transformation of STR lengths, we repeated the logistic regression test on untransformed genotypes using the same model and covariates. We also applied a non-parametric test (Mann-Whitney test) without covariates. Application of either the untransformed STR lengths in a logistic regression model or a non-parametric Mann-Whitney test led to highly similar results (correlation Pearson r^2^=0.91 and 0.94 respectively) (**Supplementary Fig. 3a-b**). We further performed the association analyses using genotypes generated by a second software program called gangSTR^20^. The STR association near *APOE* remained statistically significant using either of these alternative statistical models or using genotypes from gangSTR.

We replicated our association test in the Religious Orders Study/Memory Aging Project (ROSMAP; n=309 cases, 279 controls). The expansion at chr19:44921097-44921125:TTTA_N_ repeat near the *APOE* gene robustly replicated across these additional cohorts (meta-analysis p-value 1.43 x 10^−12^). Since this STR is near the *APOE* gene that is known to be associated with AD risk, we next assessed whether this STR represents an independent association. When re-performing the STR association test with *APOE* genotype as a covariate, the statistical association of this STR was fully attenuated (association p-value = 0.079 after adjusting for *APOE* genotype), suggesting that this STR is in linkage disequilibrium with the *APOE* genotype and does not represent an independent genetic association. Together, these results suggest that there is not a predominant STR that independently drives genetic risk for AD.

Additionally, there were 11,511 STRs nominally associated with AD risk at a p-value threshold of p<0.05. Among these nominally-associated STRs, we noticed that there was a clear bias toward more STRs having longer mean STR tract lengths in individuals with AD cases as compared to STRs having longer mean tract lengths in controls (**Fig. 2b**). This contrasts with the null hypothesis, in which we would expect an equal number of STRs with longer mean tract lengths in AD cases as there are STRs with longer mean tract length in controls. This skew toward more STRs with longer tract lengths in cases versus longer in controls was more marked at increasingly stringent p-value cutoffs. For example, at an association p-value threshold of <0.001, there were 1.15-fold as many STRs where the mean tract length was longer in cases than in controls (**Fig. 2c**). Together, these results show that while no single STR drove genetic risk of AD, longer STR tract lengths appeared to be systematically associated with higher AD risk.

In the above analysis, we treated STR tract lengths as a continuous variable for association testing. However, in known STR-associated diseases, there is usually a threshold at which a given STR tract length becomes pathogenic^9^, suggesting that large expansions of STR tract lengths rather than population variation confer disease status. Thus, we performed association analyses to test whether there is a difference in the burden of STR expansions for each STR between AD cases and controls (**Fig. 1a**, model 2). The challenge with testing this model is that the tract length at which an STR becomes pathogenic is not known *a priori*. We thus performed our analyses at different STR tract length thresholds of ≥1, ≥5, ≥10, or ≥20 repeat units longer than the GRCh38 reference genome.

For each STR, we counted the number of case and control individuals with and without an STR expansion as defined by a given STR tract length threshold and performed a hypergeometric test to assess for differences in number of expansions between case and control individuals. We found that association results were well-calibrated across all STR tract length thresholds tested (**Fig. 2d; Supplementary Table 3**). However, besides the chr19:44921097-44921125:TTTA_N_ STR near *APOE* at a STR tract length threshold of ≥1 repeat unit (p-value=7.74×10^−11^, two-tailed Fisher’s exact test), there were no other STRs that were statistically significant after correcting for multiple hypothesis testing.

While no single STR showed a statistically significant difference in the number of expansions between case and control individuals, we next tested whether there may be a difference in the cumulative burden of expansions across the genome in cases versus controls. We identified STR expansions at different STR length thresholds (≥1, ≥5, ≥10, or ≥20 repeat units longer than reference) and at different STR expansion frequency cutoffs (seen once, ≤5, ≤10, ≤100, or no cutoff in the combined n=2,981 individuals) and counted the total number of STR expansions per individual. Strikingly, we found that individuals with AD carried a higher burden of longer and rare STR expansions. For example, individuals with AD carried a 3.35-fold higher number of STRs ≥20 repeat units longer than the reference and seen only once in the cohort (p-value=3.88×10^−16^, two-tailed Mann-Whitney test) (**Fig. 2e**). This observed increased burden of STRs in individuals with AD was attenuated when examining either shorter STR alleles or more common alleles. For example, there was only a 1.10-fold increased burden in AD individuals for STRs ≥1 repeat unit longer than reference and seen once in the cohort. Similarly, there was only a 1.05-fold increased burden in AD individuals for STRs ≥20 repeat units longer than reference but when no frequency cutoff was placed. These results suggest that a high burden of rare expanded STR alleles is strongly associated with risk of AD.

### Identification of an increased burden of rare STRs expansions in AD

Given our finding of an increased burden of STR expansions in individuals with AD, we next sought to systematically identify rare, long STR expansions without pre-specifying an STR length threshold since the tract length threshold is not known *a priori* for any given STR. To increase statistical power to identify rare STR expansions, we combined samples from across the ADC and ROSMAP cohorts (total n=1,492 controls and 1,489 cases). Since we are focused on rare expansions, we lack statistical power to test for the association between rare expansions in AD case/control status. Thus, for remaining analyses, we aggregate the number of expansions per individual and test whether there is a difference in the burden of rare STR expansions between case and control individuals (**Fig. 1a**, model 3).

To detect STR expansions, we implemented an approach using density-based spatial clustering of applications with noise (DBSCAN), which we extended from the work of Trost *et al*.^13^. This approach obviates the need for selecting an arbitrary expansion length threshold and frequency cutoff, but instead identifies individuals carrying outlier STR tract lengths for each STR. In our implementation of DBSCAN, we additionally accounted for the effects of sample and technical covariates. Applying DBSCAN, we identified expansions in 9,642 unique STRs across the genome. We classified expansions to those seen only in AD cases (n=4,412 STRs), expansions seen only in controls (n=3,365 STRs), or seen in both cases and controls (n=1,865 STRs) (**Fig. 3a; Supplementary Table 4-5)**. STRs expansions observed in AD cases were slightly but statistically significantly longer than those seen in controls (mean tract length of 39.3 and 37.3 repeat units longer than the GRCh38 reference genome tract length, respectively; p<1.1×10^−3^, two-sided Mann-Whitney test) (**Fig. 3b**). 50.1% of AD STR expansions were dinucleotide repeats and just 2.1% of AD STR expansions had a repeat unit of 6 bp or longer (**Fig. 3c**). Many of the STR expansions were observed in more than one individual (**Supplementary Fig. 4**). 1,497 STR expansions were present in more than one AD case individual, including 213 STR expansions present in five or more AD case individuals.

**Figure 3:**
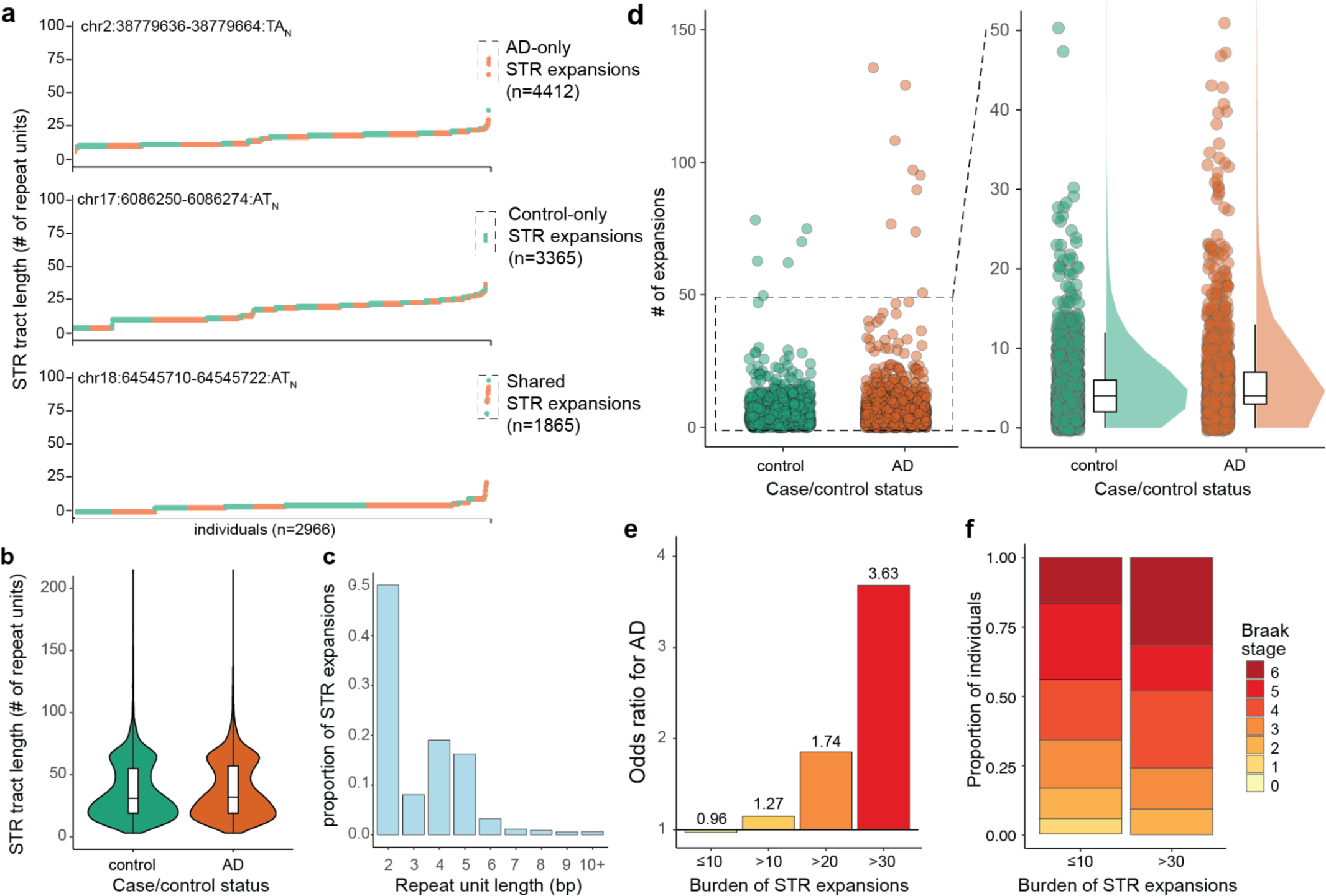
DBSCAN identifies increased burden of rare STR expansions in AD. **a,** Three separate STRs are shown as examples of AD-only STR expansions, control-only STR expansions, or shared STR expansions as identified by DBSCAN. For each STR, y-axis shows tract length for the STR, and the x-axis reflects the individuals (each represented by one point and ranked by their tract length). Orange points represent AD cases and green points represent controls. **b,** STR tract lengths in number of repeat units longer than GRCh38 reference genome for control STR expansions (green) and AD STR expansions (orange). **c,** Repeat unit lengths for STR expansions seen in controls (green) and AD cases (orange). Histogram shows values as proportion of STR expansions seen in controls and cases respectively. **d,** Number of STR expansions identified by DBSCAN per individual in controls (green) versus cases (orange). Each point represents one individual. Right panel is zoomed in for individuals with fewer than 50 STRs. **e,** Odds ratio for AD case/control status for individuals carrying varying numbers of STR expansions. Odds ratios > 1.0 represent higher odds of having AD. **f,** Stacked barplot of Braak stages for individuals with ≤ 10 expansion (top) or > 30 expansions (bottom). Braak stages are from 0-6, with higher values reflecting more severe neuropathology.

We next tested for differences in the burden of STRs in AD cases versus controls. There was a mean of 6.27 and 5.27 STR expansions in AD cases versus controls, representing a 1.19-fold higher burden of STR expansions in cases (p=8.27×10^−3^, two-sided Mann Whitney test) (**Fig. 3d**). The median number of STR expansions was 4.0 for both AD cases and controls. Strikingly, we found that individuals who carried > 30 STR expansions had an odds ratio of 3.62 for having AD (**Fig. 3e)**.

We next ascertained if our observations were reproducible using gangSTR^20^, a separate STR genotyping algorithm. We note that there were fewer STR expansions identified by gangSTR (mean 1.028, median 1.00 per individual) as compared to ExpansionHunter (mean 5.77, median 4.00 per individual). This is consistent with prior work demonstrating higher sensitivity of ExpansionHunter for identifying STR expansions as compared to gangSTR and that ExpansionHunter tends to overestimate STR tract lengths while gangSTR underestimates them^26^. Using gangSTR, we again found a higher burden of STR expansions in cases (p=2.93×10^−4^, two-sided Mann Whitney test) (**Supplementary Fig. 5a**). We also found that individuals who carry >10 STR expansions as identified by gangSTR have a 3.50 odds ratio for having AD (**Supplementary Fig. 5b**).

We next assessed whether individuals with a high burden of STR expansions also had differences in neuropathology as measured by Braak staging which reflects the degree of spread of tau pathology^28^. Braak staging data was available for 1,188 individuals of the 2,981 individuals in the cohort (n=365 controls and 823 AD cases). Braak stages are scored from 0 to 6, with 0 representing absence of AD neuropathology and 6 being the most severe spread of AD neuropathology^28^. We found that individuals with > 30 expansions had worse AD neuropathology compared to individuals with ≤10 expansions (p=0.01, Kruskal-Wallis rank sum test) (**Fig. 3f**). There was also a weak association between the number of outliers and APOE genotype (p=0.013, Kruskal Wallis rank sum test) and with participant age (p=1.88×10^−4^, Spearman correlation) (**Supplementary Fig. 6a-b**).

### STR expansions are enriched in active regulatory regions of the genome

The majority of STRs tested in our study are not within coding portions of genes (**Fig. 1b**), precluding any simple interpretations of the mechanisms by which they may promote disease pathogenesis. We first annotated the genomic distributions of the 6,276 AD STR expansions and found that they had similar distributions to the background of 293,763 STRs we tested (see Methods). Specifically, the majority of AD STR expansions were in distal intergenic regions (38.1%; >3kb from the nearest TSS) or in promoter regions (14.7%; defined as being ≤3 kb upstream from the nearest TSS) (**Fig. 4a**). However, AD STR expansions (median 24.7 kb) were further from the nearest TSS than background STRs (median 22.2 kb) (p-value=6.10×10^−10^, two-sided Student’s t-test), which appeared to be driven by a slightly larger subset of STR expansions that were >100 kb upstream of the nearest TSS (**Fig. 4b**).

**Figure 4.**
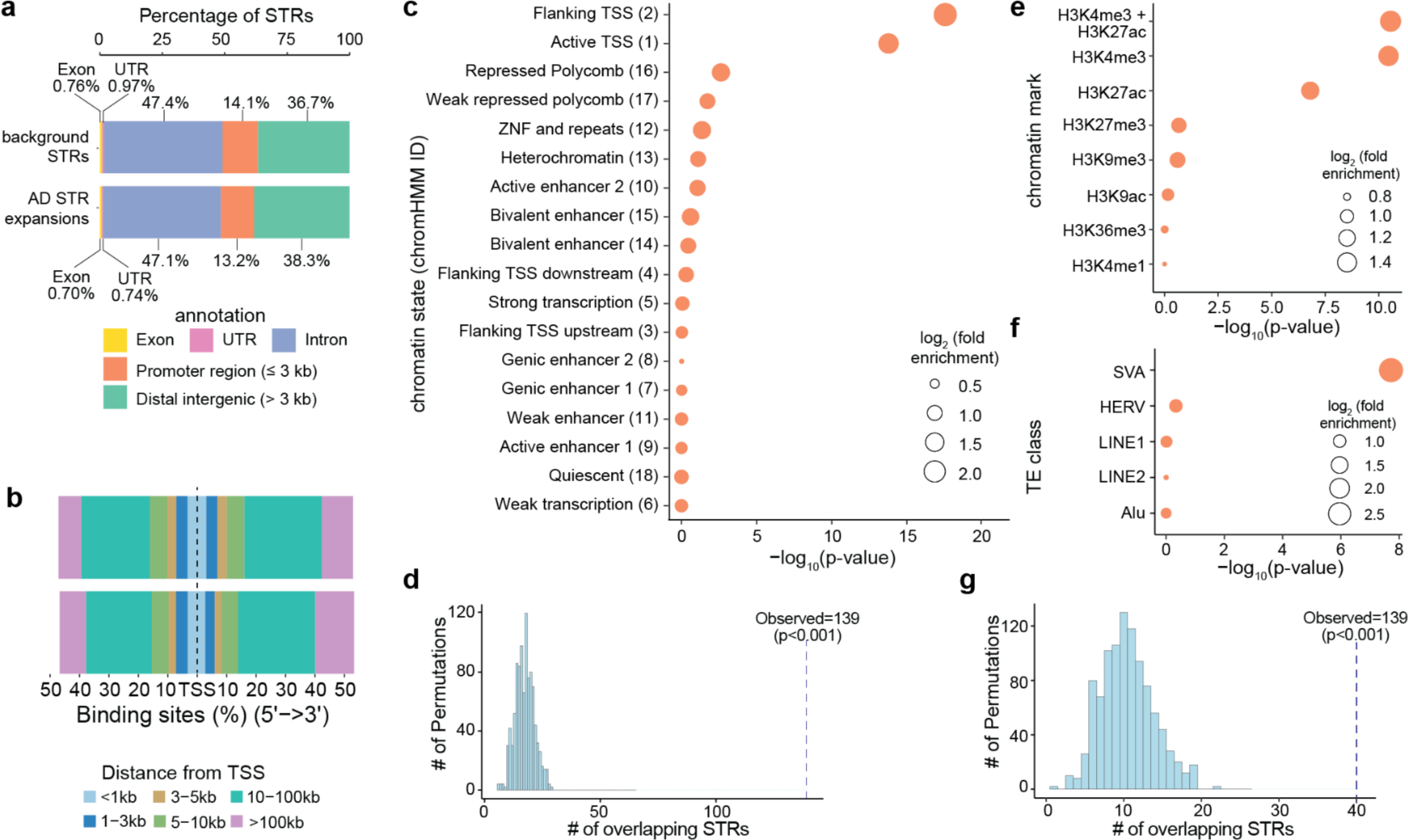
STR expansions in AD are enriched in active promoters and SVA transposable elements. **a.** Genomic distribution of all (background) STRs tested (top) compared to AD STRs expansions (bottom). **b**, Distribution of distances to nearest TSS for all STRs tested (top) or AD STR expansions (bottom). **c,** Enrichment of AD STR expansions in different chromatin states from the 18-state chromHMM model for adult hippocampus. **d,** Enrichment of AD STR expansions Dotted vertical line shows the number of AD STR expansions observed to overlap chromatin state 2 in hippocampus. in “flanking TSS” (chromatin state 2 from chromHMM) in adult hippocampus using a permutation-based test. **e,** Enrichment of AD expanded STRs in peaks from different histone marks based on ChIP-seq in bulk adult hippocampal tissue. **f,** Enrichment of AD STR expansions in different TE classes. **g,** Enrichment of AD expanded STRs in SVA elements using a permutation-based test. For **c, e,** and **f,** height of points along the x-axis represents the statistical significance of enrichment, as measured by two-sided Fisher’s exact test. Size of points represents the magnitude of enrichment as represented by the log_2_(fold enrichment), and points are ordered by statistical significance for AD STR expansions. For **d** and **g,** dotted line reflects the observed number of AD STR expansions overlapping the genomic annotation. Histogram represents the number of AD STR expansions overlapping each of 1000 randomly permuted genomic regions.

Given that the majority of AD STR expansions were not within protein-coding regions, we next explored whether these AD STRs may be enriched in any chromatin features. We tested whether AD STR expansions were enriched in chromatin states based on bulk post-mortem hippocampus ChIP-seq data from ENCODE^29^, using the 18 chromatin state partitions in chromHMM^30^. We found that “flanking TSS” (chromatin state 2) and “active TSS” (chromatin state 1) reflecting active promoter regions were the most enriched chromatin states for AD STR expansions (p-value=2.82×10^−18^ and 1.66×10^−14^ respectively, two-tailed Fisher’s exact test) (**Fig. 4c**). We validated the enrichment using a permutation-based approach by randomly drawing genomic regions and testing for their overlap with AD STR expansions. This permutation-based approach recapitulated the enrichment of AD STR expansions in “flanking TSS” (empirical enrichment p-value < 0.001) (**Fig. 4d**). The enrichment of AD STR expansions in “flanking TSS” was stronger in brain tissues compared to tissues from other parts of the body (**Supplementary Fig. 7**). We also validated the chromHMM enrichment results using ChIP-seq data from post-mortem hippocampal tissue in ENCODE. Across ChIP-seq peaks for seven different histone marks, we found that AD STR expansions were most highly enriched in H3K4me3 (p=3.54 ×10^−11^, two-tailed Fisher’s exact test) and H3K27ac peaks (p=1.59×10^−7^). H3K4me3 marks active promoters, and H3K27ac marks active promoters and/or enhancers. Chromatin marked by both H3K4me3 and H3K27ac had even stronger enrichment (p=2.84×10^−11^) (**Fig. 4e**). These findings demonstrate that AD STR expansions are enriched in active promoter regions in the brain.

Given that many transposable elements (TE) contain or are in proximity to STRs^31^, we next sought to understand whether AD STR expansions are enriched for any specific TE classes. We tested for enrichment across 5 broad categories of TEs: Alu, Human Endogenous Retroviruses (HERVs), long interspersed nuclear elements (LINE)-1 and −2, and SINE-VNTR-Alus (SVA). AD STR expansions were highly enriched for SVA elements (p-value 1.90×10^−8^, two-tailed Fisher’s exact test) (**Fig. 4f**), which we also validated using a permutation-based approach (**Fig. 4g**). Our observation of a strong enrichment of STR expansions within active promoter regions in the brain is consistent with prior work suggesting that SVA elements have being co-opted for enhancers and promoters in neurons^32–34^.

### AD STR expansions are enriched in disease pathways with relevance to AD

While the majority of AD STR expansions were not within protein-coding regions (**Fig. 4a**), we sought to understand whether they may be enriched near genes with certain biological or molecular functions. For each STR expansion, we identified all genes with TSS within +/− 250 kb. We then performed a gene ontology enrichment analysis for genes near AD STR expansions (n=6276) compared to all STRs tested (n=293,763). We found that many of the most strongly associated gene sets were related to neuron biology, such as “neuron projection morphogenesis”, and “axon development” (**Fig. 5a**). Thus, while most AD STR expansions were not protein-coding, they are highly enriched near genes implicated in biological processes with known relevance to AD pathophysiology.

**Figure 5:**
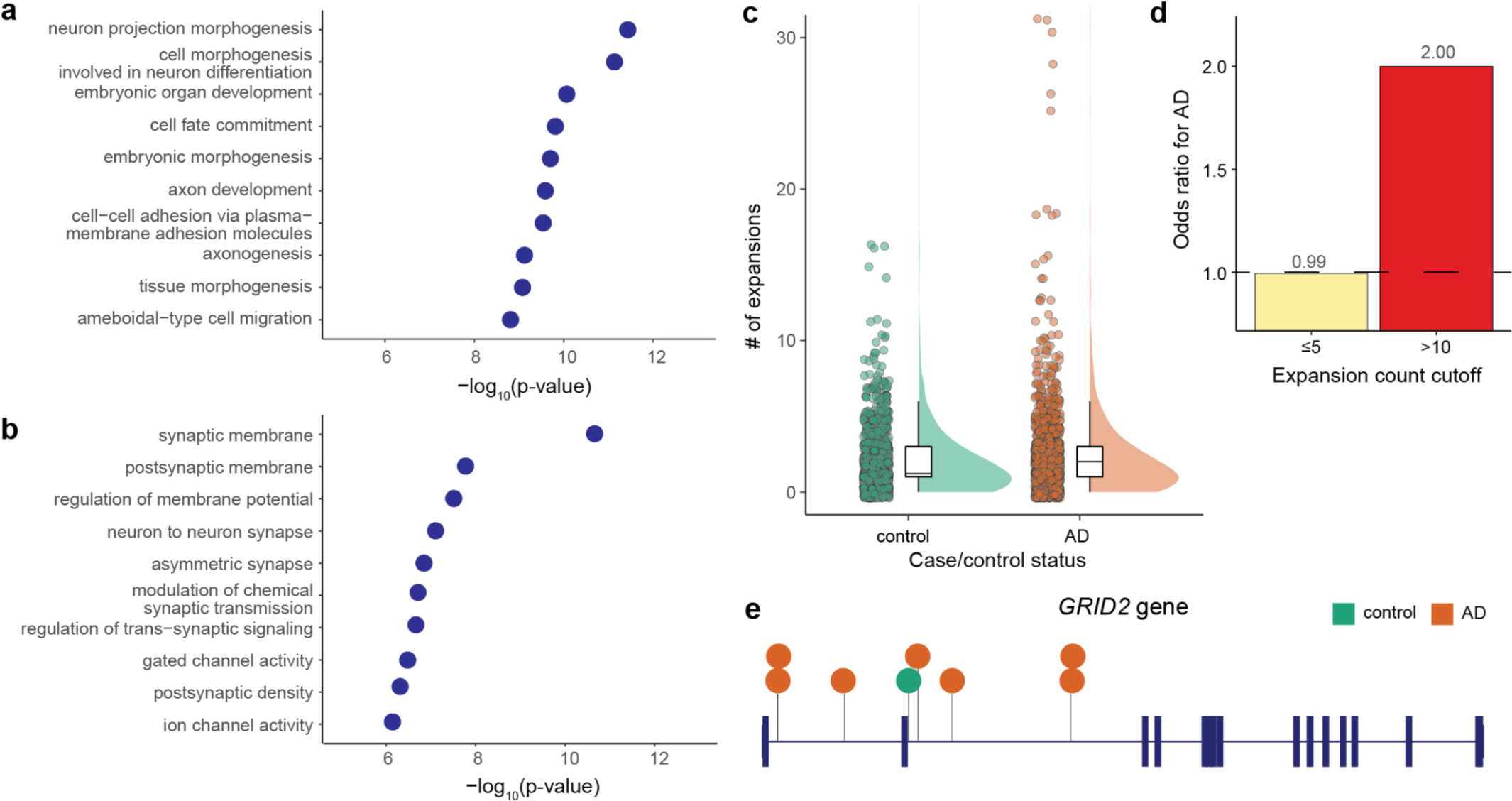
Insights into genes and pathways implicated by AD STR expansions. **a,** Gene set enrichment for AD STR expansions relative to all STRs tested. The top 10 most highly associated gene ontology terms are shown. Height of points along the x-axis represents the statistical significance of enrichment, as measured by two-sided Fisher’s exact test. **b,** Same as a, except only STRs expansions within gene bodies. **c,** Number of STR expansions within gene bodies identified by DBSCAN per individual in controls (green) versus cases (orange). **d,** Odds ratio for AD case/control status for individuals carrying varying numbers of STR expansions within gene bodies. Odds ratios > 1.0 represent higher odds of having AD. **e,** *GRID2* gene is shown with thick vertical lines representing exons and horizontal lines representing introns. Locations of AD STR expansions (orange) and control individual STR expansions (green) are shown. Each circle represents an STR expansion seen in one individual.

Since assigning intergenic STR expansions to a cognate gene is challenging, we next examined AD STR expansions in gene bodies (i.e., those occurring in exons, untranslated regions, or introns of genes). We compared AD STR expansions found within gene bodies (n=1,327) to all STRs tested within gene bodies (n=13,594) and identified strong enrichments in many gene sets related to synaptic function (**Fig. 5b**). These enrichments are notable given the pathologic role of synaptic dysfunction in AD^35^. We also found a higher burden of STR expansions in gene bodies between AD cases and control individuals (p=0.021, two-sided Mann-Whitney test) (**Fig. 5c**). Moreover, individuals with more than 10 STR expansions in gene bodies had a 2.0 odds ratio for having AD (**Fig. 5d**). There was no difference in the number of STR expansions in protein-coding regions only between cases and controls (p=0.89, two-sided Mann-Whitney test), though the number of such expansions was low (median 0 and mean 0.20 protein-coding STR expansions per individual).

As an example, we highlight the *GRID2* gene, in which we identified seven STR expansions across nine individuals with AD compared to just one STR expansion seen in a single control individual (**Fig. 5e**). *GRID2* encodes a subunit of the glutamate receptor and has recognized roles in synaptic transmission^36^. Together, these results demonstrate that AD STR expansions are highly enriched in and near genes implicated in biological processes with known relevance to AD pathophysiology and thus the AD STR expansions may represent molecular drivers of disease. Moreover, these results show that while these STR expansions were detected in blood-derived DNA, they appear to function in the brain, where much of the pathophysiology of AD is presumed to occur.

## Discussion

In this work, we performed genome-wide profiling of STRs in 2,981 with and without AD to understand whether STRs associate with risk of AD. In contrast to the known STR-associated disorders, we did not find that a single STR individually drove genetic risk for AD. Instead, we identified thousands of STR expansions distributed throughout the genome with a higher burden of STR expansions in cases as compared to controls. Moreover, expanded STRs in AD cases were enriched in active promoters in the brain and in SVA TEs. These results suggest a model in which a polygenic burden of STR expansions distributed throughout the genome promotes risk of AD.

Remarkably, we found that individuals with AD carried an excess burden of expanded STRs. Each individual with AD had a mean of 6.27 STR expansions, though we note this estimate is likely an underestimate given the lower sensitivity of STR genotyping software on long expansions and our stringent quality filters. Nonetheless, this suggests that a polygenic burden of expanded STRs rather than a single STR promotes risk of AD. This polygenic effect is similar to what has been observed for STRs in autism spectrum disorder^11,13^ and in schizophrenia^12^. The distributed nature of these AD-associated STR expansions throughout the genome rather than within one or a few genes suggests that general genomic instability is a pathologic hallmark of the genomes in AD. This concept of an increased burden of STRs in individuals with AD is consistent with prior findings that individuals with AD have a higher burden of rare coding single nucleotide variants^37^ and structural variants^38^.

This observation of an increased burden of STR expansions in AD suggests one of two mechanistic models, which are not mutually exclusive. First, STR expansions may be an epiphenomenon of a disease process that promotes genomic instability. For example, AD pathology or biological aging may promote STR instability to result in an increased number of STR expansions in AD. The second possibility is that inherited and/or somatically unstable STR expansions promote disease pathogenesis. In support of this second model, we find that STR expansions in cases are preferentially localized in active promoters of genes with important neuronal functions. These results suggest that STR expansions may have a functional role in AD onset and progression, though mechanistic studies will be needed to dissect the exact causal relationships between STRs and AD.

Strikingly, we found that AD-associated STR expansions were colocalized within active promoters in brain tissues. The AD STR expansions were particularly enriched within SVA elements, which have been proposed to have been co-opted for enhancers and promoters during human evolution^32,33,39^. SVA elements have also been shown to be important sources of tandem repeat variation and evolution in the human genome and particularly near neuronal genes^34^. Thus, our findings suggest that STRs at these SVA elements in active gene regulatory elements are prone to instability, particularly near genes with roles in AD pathophysiology. These findings in the context of existing literature underscore an important relationship between TEs, genome instability, and disease.

There are several important limitations to our study. First, while we expect molecular drivers of AD to act in the brain, we only have access to peripheral blood-derived DNA. We did however find that AD STR expansions were enriched in active promoters in the brain and near genes involved in neuronal function, suggesting that even though STR lengths were measured in the blood, they may manifest their effect on AD risk in the brain. Similarly, without paired blood and brain-derived DNA, we were unable to test for somatic instability of STRs in brain tissues. Second, we used short-read WGS where the accuracy of genotyping STRs is limited, particularly for longer STR alleles^26^. We mitigated these concerns by using two different software programs to replicate our results. We also compared our STR lengths to well-matched control individuals; as such, we would expect that any STR genotyping artifacts would be randomly allocated between AD and control individuals. Finally, our study design uses observational data in humans and so we cannot directly test causality. Future studies performing *in vitro* mechanistic dissection of these STRs will be required to establish a causal role of STRs in AD.

Together, our work identifies novel links between STR expansions and regulatory elements in AD. These results underscore the importance of uncovering the role of STRs in a broad range of diseases and understanding the mechanisms by which these STRs may promote disease risk.

## Methods

### Cohort Description

The Alzheimer’s Disease Sequencing Project (ADSP) is a collaborative project aiming at identifying new variants, genes, and therapeutic targets in AD^40^. In the R3 release of ADSP, genetic data was collected from 16,905 individuals were collected across 24 cohorts and whole genome sequencing was performed by Illumina HiSeqX, HiSeq2000, HiSeq2500, and NovaSeq platforms.

In this work, we used individuals from the ADC and ROSMAP cohorts within the ADSP. We utilized AD case/control status as adjudicated and provided by the ADSP. We restricted our analyses to individuals with self-reported non-Hispanic White ethnicity. We further restricted to individuals with European genetic ancestry based on PCA, which were provided by the ADSP. Based on manual inspection of PCA plots, We retained samples with PC1< −0.0037 and PC2 < 0.02. To minimize the impact of sequencing coverage on STR calls, we removed samples with < 30X or > 50X sequncing coverage across the genome.

### Generation of polymorphic STR panel

To reduce computational burden, we generated a custom panel of polymorphic STRs for testing rather than testing all STRs genome-wide. To generate this panel, we first ran gangSTR^20^. using the hg38_ver13 catalog provided by the gangSTR authors (https://s3.amazonaws.com/gangstr/hg38/genomewide/hg38_ver13.bed.gz) on 246 AD cases and 249 AD controls from the NIA ADC cohort. We used gangSTR v2.4.0 on this hg38_ver13 reference panel using default parameters, except --max-proc-read 100000 and –rescue-count 10. STRs were determined to be polymorphic if at least one individual in the cohort had a non-reference STR length and the genotyping rate across the 495 samples was ≥ 90%. In total, there were 237,197 STRs that met these criteria. We next merged these 237,197 STRs with a set 174,262 STRs previously identified to be polymorphic^22^. This resulted in a union set of 321,742 unique polymorphic STRs.

### Genotyping STR tract lengths

We genotyped STR tract lengths using ExpansionHunter v5^22^ on our panel of 321,742 polymorphic STRs using default parameters. Prior to running ExpansionHunter, the polymorphic STR panel was converted to json format required for ExpansionHunter using custom python scripts.

We also genotyped STR tract lengths using gangSTR^20^ on our panel of 321,742 polymorphic STRs. To increase sensitivity of gangSTR, we added in offtarget alignment locations for any STR that had a p-value < 0.05 in the ExpansionHunter single STR association analysis. To determine offtarget alignment locations, we used wgsim v1.11 to simulate 10000 sequencing reads for each STR (https://github.com/lh3/wgsim). We used the following parameters in wgsim: -e 0.005 -d 500 -s 100 -N 10000 −1 150 −2 150 -r 0 -R 0 -X 0. Simulated reads were then realigned back to the GRCh38 reference genome using bwa mem version 0.7.17 with default parameters^41^. The offtarget alignment locations of simulated reads in the GRCh38 reference genome were then extracted using scripts provided by the gangSTR authors, and we used the top 5 offtarget locations for each STR. gangSTR v2.4.2 was run on this custom reference panel with default parameters except --max-proc-read 100000.

### Single STR association analysis

We performed association analyses of STR genotype with AD risk under a dominant model by testing the longer of two alleles for each STR. Given that many of the STR genotypes were not normally distributed, we first performed rank-based inverse normal transformation of STR genotypes. To test for the association of each STR with AD case/control status, we applied a logistic regression model, controlling for sample covariates (sex, age, and the first three PCs) and technical covariates (genomic sequencing coverage for each sample, local sequencing coverage for each sample at the STR, and sequencing center).

We report the logistic regression p-value of the rank-based inverse normal transformed STR genotype on AD risk. We separately report the regression p-value in a logistic regression model without inverse normal transformation to derive absolute effect sizes on AD case/control status. Finally, to ensure results were not due to the non-parametric nature of the STR genotypes, association analyses were also performed using a non-parametric association test (two-sided Mann-Whitney test) without covariates.

For reporting of STR association analyses, we removed all STRs that were in segmentally duplicated regions (http://hgdownload.cse.ucsc.edu/goldenpath/hg38/database/genomicSuperDups.txt.gz) (n=27,617), resulting in a final list of 293,785 STRs that were reported in our association analyses. Multiple hypothesis testing correction was performed using Bonferroni correction, with a p-value threshold of 1.70×10^−7^ (0.05/293785).

### Hypergeometric test for STR expansions

We performed a burden test for the number of STR expansions in case versus control individuals for each STR. We performed this analysis using ExpansionHunter genotyping data on the ADC + ROSMAP cohort.

We first identified STR expansions for each of the 293,785 STRs in our custom STR panel not located within segmental duplications. We identified STR expansions as STR tract lengths that were ≥ 1, 5, 10, or 20 repeat units longer than the GRCh38 reference STR tract length. For each STR, we then constructed a 2×2 contingency table of the number of individuals with and without an STR expansion as defined by these thresholds. We applied a two-sided Fisher’s exact test to each STR to test whether there is a difference in burden of STR expansions in cases versus controls. Multiple hypothesis testing correction was performed using Bonferroni correction, with a p-value threshold of 1.70×10^−7^ (0.05/293785).

We also performed analyses restricting to expansions observed in only one individual, ≤5, ≤10, ≤100, or with no frequency cutoff in the combined n=2,981 individuals. We then compared the total number of STR expansions in AD case versus control individuals using a two-sided Mann-Whitney test.

### Identifying STR expansions using DBSCAN

We applied the DBSCAN outlier detection method to identify STR expansions, which we extended from the work of Trost et al.^13^. Briefly, DBSCAN is an unsupervised density-based clustering method that can be used to identify outlier groups^42^, here representing extreme STR tract lengths for each STR. DBSCAN defines a cluster based on the minimum number of data points (**μ**) reachable to each other by a maximum distance (**ε**). Data points not reachable by the clusters are classified outliers if they have an STR tract length that is higher than those of cluster members. Specifically, **ε** was set as the maximum of 2 x mode of STR lengths, and **μ** was set as the log_2_ of the number of samples.

For each STR, the longer of the two alleles for each individual was used as input for DBSCAN. To account for potential confounders, we first performed linear regression for the tract lengths of each STR to regress out the effects of sample covariates (sex, age, and the first three PCAs) and technical covariates (genomic sequencing coverage for each sample, local sequencing coverage for each sample at the STR, and sequencing center). We input the residuals from the linear regression into DBSCAN, with **ε** and **μ** as determined above. We ran DBSCAN on these residuals to identify outlier STR lengths. We applied DBSCAN separately for STR genotyping results from ExpansionHunter and gangSTR. DBSCAN was implemented using the dbscan package v1.1-11 (https://github.com/cran/dbscan) in R v3.6.3^43,44^.

### Testing for enrichment of STRs in chromHMM and ChIP-seq annotations

We downloaded chromHMM partitions under the 18 state model for the adult human hippocampus (ENCODE tissue ID E071) from the Roadmap Epigenomics Consortium (https://egg2.wustl.edu/roadmap/web_portal/chr_state_learning.html)^29,30^. We then tested whether AD STR expansions were enriched in each chromatin state, relative to a background of all STRs in our custom panel of polymorphic STRs. We performed statistical testing using a two-sided Fisher’s exact test. Prior to statistical testing, we removed all STRs located within segmental duplications from the foreground and background STR sets. We considered an STR to be located within a chromHMM partition if at least one bp of the STR was within the partition.

We also tested whether AD STR expansions were enriched in certain histone marks as assayed by ChIP-seq. We downloaded ChIP-seq peak data from ENCODE for the adult hippocampus for H3K4me1, H3K4me3, H3K9ac, H3K27ac, H3K27me3, and H3K36me3. For each biological sample, we used IDR-replicated peaks from ENCODE. If there were multiple biological samples for each histone mark, we merged peaks across samples using BEDTools v2.26^45^. We then performed the enrichment analysis as above.

For both the chromHMM data and ENCODE ChIP-seq data, we also performed enrichment analyses using a permutation-based approach. We calculated how many of the n=6276 AD STR expansions directly overlapping the peak set. We then generated 1000 random sets of 6276 peaks using regioneR v1.30.0 using default parameters except per.chromsome=F^46^. Random peak sets were generated against the GRCh38 reference genome, with masking of segmental duplications (see above). We counted how many AD STR expansions directly overlapped each random peak set. We derived an empirical p-value by counting the number of random permutations with equal or more overlaps than the observed number of overlaps and dividing by 1000.

### Testing for STR enrichments in transposable elements

To identify locations of TE in the genome, we obtained the RepeatMasker file from the UCSC Table Browser (https://genome.ucsc.edu/cgi-bin/hgTables)^47^. We tested for enrichment of AD STR expansions across 5 broad classes of TEs: Alu, HERVs, LINE1, LINE2, and SVA elements. For each TE class, we tested whether AD STR expansions were enriched in the TE, relative to a background of all STRs in our custom STR panel. We performed statistical testing using a two-sided Fisher’s exact test. Prior to statistical testing, we removed all STRs located within segmental duplications from the foreground and background STR sets. We considered an STR to be located within a TE if at least one bp of the STR was within the TE coordinates from the RepeatMasker file.

### Gene Ontology Enrichment Analysis

We performed gene set enrichment analysis using the clusterProfiler v4.2.2 package in R v3.6.3^48^. For all STRs, we compared AD STR expansions (n=6276) to all STRs in the reference STR panel (n=293,763). For this analysis, we assigned each STR to the gene with the closest TSS within 500 kb. We also performed separate analyses for STRs within the gene body (either introns, exons, or untranslated regions), where we compared genes with AD STR expansions within the gene body (n=1327) to all genes with STR from the reference STR panel present (n=13,594). We then tested the enrichment of genes assigned to AD STR expansions compared to genes assigned to background STRs using the enrichGO function in clusterProfiler, with the following parameters: keyType="ENTREZID", ont=”ALL”, pvalueCutoff = 0.05, qvalueCutoff = 0.05.

## Data Availability Statement

The data analyzed in this study is subject to the following licenses/restrictions: Data is accessible from NIAGADS DSS *via* qualified access. Formal requests to access these datasets can be submitted to NIAGADS DSS: https://dss.niagads.org/. All code used in this manuscript is available at https://github.com/mhguo1/AD_STR.

## Data Availability

The data analyzed in this study is subject to the following licenses/restrictions: Data is accessible from NIAGADS DSS via qualified access. Formal requests to access these datasets can be submitted to NIAGADS DSS: https://dss.niagads.org/.

## Acknowledgements

We thank members of the Cremins lab for helpful discussions. This work is supported by NIH National Institute of Neurologic Disorders and Stroke (R25 NS065745 to MHG), University of Pennsylvania Christopher Clark Scholars Award (to MHG), NIH National Institute on Aging (RF1-AG074328, P30-AG072979, U54-AG052427, and U24-AG041689 to WPL), NIH National Institute of Mental Health (1R01MH120269 to JEPC), NSF CAREER Award (CBE-1943945 to JEPC), NSF EFRI Award (EFMA1933400 to JEPC), and Chan Zuckerberg Initiative Neurodegenerative Disease Pairs Awards (2020-221479-5022 and DAF2022-250430 to JEPC).

Data for this study were prepared, archived, and distributed by the National Institute on Aging Alzheimer’s Disease Data Storage Site (NIAGADS) at the University of Pennsylvania (U24-AG041689), funded by the National Institute on Aging.

## Alzheimer’s Disease Sequencing Project

The Alzheimer’s Disease Sequencing Project (ADSP) is comprised of two Alzheimer’s Disease (AD) genetics consortia and three National Human Genome Research Institute (NHGRI) funded Large Scale Sequencing and Analysis Centers (LSAC). The two AD genetics consortia are the Alzheimer’s Disease Genetics Consortium (ADGC) funded by NIA (U01 AG032984), and the Cohorts for Heart and Aging Research in Genomic Epidemiology (CHARGE) funded by NIA (R01 AG033193), the National Heart, Lung, and Blood Institute (NHLBI), other National Institute of Health (NIH) institutes and other foreign governmental and non-governmental organizations. The Discovery Phase analysis of sequence data is supported through UF1AG047133 (to Drs. Schellenberg, Farrer, Pericak-Vance, Mayeux, and Haines); U01AG049505 to Dr. Seshadri; U01AG049506 to Dr. Boerwinkle; U01AG049507 to Dr. Wijsman; and U01AG049508 to Dr. Goate and the Discovery Extension Phase analysis is supported through U01AG052411 to Dr. Goate, U01AG052410 to Dr. Pericak-Vance and U01 AG052409 to Drs. Seshadri and Fornage.

Sequencing for the Follow Up Study (FUS) is supported through U01AG057659 (to Drs. PericakVance, Mayeux, and Vardarajan) and U01AG062943 (to Drs. Pericak-Vance and Mayeux). Data generation and harmonization in the Follow-up Phase is supported by U54AG052427 (to Drs. Schellenberg and Wang). The FUS Phase analysis of sequence data is supported through U01AG058589 (to Drs. Destefano, Boerwinkle, De Jager, Fornage, Seshadri, and Wijsman), U01AG058654 (to Drs. Haines, Bush, Farrer, Martin, and Pericak-Vance), U01AG058635 (to Dr. Goate), RF1AG058066 (to Drs. Haines, Pericak-Vance, and Scott), RF1AG057519 (to Drs. Farrer and Jun), R01AG048927 (to Dr. Farrer), and RF1AG054074 (to Drs. Pericak-Vance and Beecham).

The ADGC cohorts include: Adult Changes in Thought (ACT) (U01 AG006781, U19 AG066567), the Alzheimer’s Disease Research Centers (ADRC) (P30 AG062429, P30 AG066468, P30 AG062421, P30 AG066509, P30 AG066514, P30 AG066530, P30 AG066507, P30 AG066444, P30 AG066518, P30 AG066512, P30 AG066462, P30 AG072979, P30 AG072972, P30 AG072976, P30 AG072975, P30 AG072978, P30 AG072977, P30 AG066519, P30 AG062677, P30 AG079280, P30 AG062422, P30 AG066511, P30 AG072946, P30 AG062715, P30 AG072973, P30 AG066506, P30 AG066508, P30 AG066515, P30 AG072947, P30 AG072931, P30 AG066546, P20 AG068024, P20 AG068053, P20 AG068077, P20 AG068082, P30 AG072958, P30 AG072959), the Chicago Health and Aging Project (CHAP) (R01 AG11101, RC4 AG039085, K23 AG030944), Indiana Memory and Aging Study (IMAS) (R01 AG019771), Indianapolis Ibadan (R01 AG009956, P30 AG010133), the Memory and Aging Project (MAP) (R01 AG17917), Mayo Clinic (MAYO) (R01 AG032990, U01 AG046139, R01 NS080820, RF1 AG051504, P50 AG016574), Mayo Parkinson’s Disease controls (NS039764, NS071674, 5RC2HG005605), University of Miami (R01 AG027944, R01 AG028786, R01 AG019085, IIRG09133827, A2011048), the Multi-Institutional Research in Alzheimer’s Genetic Epidemiology Study (MIRAGE) (R01 AG09029, R01 AG025259), the National Centralized Repository for Alzheimer’s Disease and Related Dementias (NCRAD) (U24 AG021886), the National Institute on Aging Late Onset Alzheimer’s Disease Family Study (NIA-LOAD) (U24 AG056270), the Religious Orders Study (ROS) (P30 AG10161, R01 AG15819), the Texas Alzheimer’s Research and Care Consortium (TARCC) (funded by the Darrell K Royal Texas Alzheimer’s Initiative), Vanderbilt University/Case Western Reserve University (VAN/CWRU) (R01 AG019757, R01 AG021547, R01 AG027944, R01 AG028786, P01 NS026630, and Alzheimer’s Association), the Washington Heights-Inwood Columbia Aging Project (WHICAP) (RF1 AG054023), the University of Washington Families (VA Research Merit Grant, NIA: P50AG005136, R01AG041797, NINDS: R01NS069719), the Columbia University Hispanic Estudio Familiar de Influencia Genetica de Alzheimer (EFIGA) (RF1 AG015473), the University of Toronto (UT) (funded by Wellcome Trust, Medical Research Council, Canadian Institutes of Health Research), and Genetic Differences (GD) (R01 AG007584). The CHARGE cohorts are supported in part by National Heart, Lung, and Blood Institute (NHLBI) infrastructure grant HL105756 (Psaty), RC2HL102419 (Boerwinkle) and the neurology working group is supported by the National Institute on Aging (NIA) R01 grant AG033193.

The CHARGE cohorts participating in the ADSP include the following: Austrian Stroke Prevention Study (ASPS), ASPS-Family study, and the Prospective Dementia Registry-Austria (ASPS/PRODEM-Aus), the Atherosclerosis Risk in Communities (ARIC) Study, the Cardiovascular Health Study (CHS), the Erasmus Rucphen Family Study (ERF), the Framingham Heart Study (FHS), and the Rotterdam Study (RS). ASPS is funded by the Austrian Science Fond (FWF) grant number P20545-P05 and P13180 and the Medical University of Graz. The ASPS-Fam is funded by the Austrian Science Fund (FWF) project I904), the EU Joint Programme – Neurodegenerative Disease Research (JPND) in frame of the BRIDGET project (Austria, Ministry of Science) and the Medical University of Graz and the Steiermärkische Krankenanstalten Gesellschaft. PRODEM-Austria is supported by the Austrian Research Promotion agency (FFG) (Project No. 827462) and by the Austrian National Bank (Anniversary Fund, project 15435. ARIC research is carried out as a collaborative study supported by NHLBI contracts (HHSN268201100005C, HHSN268201100006C, HHSN268201100007C, HHSN268201100008C, HHSN268201100009C, HHSN268201100010C, HHSN268201100011C, and HHSN268201100012C). Neurocognitive data in ARIC is collected by U01 2U01HL096812, 2U01HL096814, 2U01HL096899, 2U01HL096902, 2U01HL096917 from the NIH (NHLBI, NINDS, NIA and NIDCD), and with previous brain MRI examinations funded by R01-HL70825 from the NHLBI. CHS research was supported by contracts HHSN268201200036C, HHSN268200800007C, N01HC55222, N01HC85079, N01HC85080, N01HC85081, N01HC85082, N01HC85083, N01HC85086, and grants U01HL080295 and U01HL130114 from the NHLBI with additional contribution from the National Institute of Neurological Disorders and Stroke (NINDS). Additional support was provided by R01AG023629, R01AG15928, and R01AG20098 from the NIA. FHS research is supported by NHLBI contracts N01-HC-25195 and HHSN268201500001I. This study was also supported by additional grants from the NIA (R01s AG054076, AG049607 and AG033040 and NINDS (R01 NS017950). The ERF study as a part of EUROSPAN (European Special Populations Research Network) was supported by European Commission FP6 STRP grant number 018947 (LSHG-CT-2006-01947) and also received funding from the European Community’s Seventh Framework Programme (FP7/2007-2013)/grant agreement HEALTH-F4-2007-201413 by the European Commission under the programme “Quality of Life and Management of the Living Resources” of 5th Framework Programme (no. QLG2-CT-2002-01254). High-throughput analysis of the ERF data was supported by a joint grant from the Netherlands Organization for Scientific Research and the Russian Foundation for Basic Research (NWO-RFBR 047.017.043). The Rotterdam Study is funded by Erasmus Medical Center and Erasmus University, Rotterdam, the Netherlands Organization for Health Research and Development (ZonMw), the Research Institute for Diseases in the Elderly (RIDE), the Ministry of Education, Culture and Science, the Ministry for Health, Welfare and Sports, the European Commission (DG XII), and the municipality of Rotterdam. Genetic data sets are also supported by the Netherlands Organization of Scientific Research NWO Investments (175.010.2005.011, 911-03-012), the Genetic Laboratory of the Department of Internal Medicine, Erasmus MC, the Research Institute for Diseases in the Elderly (014-93-015; RIDE2), and the Netherlands Genomics Initiative (NGI)/Netherlands Organization for Scientific Research (NWO) Netherlands Consortium for Healthy Aging (NCHA), project 050-060-810. All studies are grateful to their participants, faculty and staff. The content of these manuscripts is solely the responsibility of the authors and does not necessarily represent the official views of the National Institutes of Health or the U.S. Department of Health and Human Services.

The FUS cohorts include: the Alzheimer’s Disease Research Centers (ADRC) (P30 AG062429, P30 AG066468, P30 AG062421, P30 AG066509, P30 AG066514, P30 AG066530, P30 AG066507, P30 AG066444, P30 AG066518, P30 AG066512, P30 AG066462, P30 AG072979, P30 AG072972, P30 AG072976, P30 AG072975, P30 AG072978, P30 AG072977, P30 AG066519, P30 AG062677, P30 AG079280, P30 AG062422, P30 AG066511, P30 AG072946, P30 AG062715, P30 AG072973, P30 AG066506, P30 AG066508, P30 AG066515, P30 AG072947, P30 AG072931, P30 AG066546, P20 AG068024, P20 AG068053, P20 AG068077, P20 AG068082, P30 AG072958, P30 AG072959), Alzheimer’s Disease Neuroimaging Initiative (ADNI) (U19AG024904), Amish Protective Variant Study (RF1AG058066), Cache County Study (R01AG11380, R01AG031272, R01AG21136, RF1AG054052), Case Western Reserve University Brain Bank (CWRUBB) (P50AG008012), Case Western Reserve University Rapid Decline (CWRURD) (RF1AG058267, NU38CK000480), CubanAmerican Alzheimer’s Disease Initiative (CuAADI) (3U01AG052410), Estudio Familiar de Influencia Genetica en Alzheimer (EFIGA) (5R37AG015473, RF1AG015473, R56AG051876), Genetic and Environmental Risk Factors for Alzheimer Disease Among African Americans Study (GenerAAtions) (2R01AG09029, R01AG025259, 2R01AG048927), Gwangju Alzheimer and Related Dementias Study (GARD) (U01AG062602), Hillblom Aging Network (2014-A-004-NET, R01AG032289, R01AG048234), Hussman Institute for Human Genomics Brain Bank (HIHGBB) (R01AG027944, Alzheimer’s Association “Identification of Rare Variants in Alzheimer Disease”), Ibadan Study of Aging (IBADAN) (5R01AG009956), Longevity Genes Project (LGP) and LonGenity (R01AG042188, R01AG044829, R01AG046949, R01AG057909, R01AG061155, P30AG038072), Mexican Health and Aging Study (MHAS) (R01AG018016), Multi-Institutional Research in Alzheimer’s Genetic Epidemiology (MIRAGE) (2R01AG09029, R01AG025259, 2R01AG048927), Northern Manhattan Study (NOMAS) (R01NS29993), Peru Alzheimer’s Disease Initiative (PeADI) (RF1AG054074), Puerto Rican 1066 (PR1066) (Wellcome Trust (GR066133/GR080002), European Research Council (340755)), Puerto Rican Alzheimer Disease Initiative (PRADI) (RF1AG054074), Reasons for Geographic and Racial Differences in Stroke (REGARDS) (U01NS041588), Research in African American Alzheimer Disease Initiative (REAAADI) (U01AG052410), the Religious Orders Study (ROS) (P30 AG10161, P30 AG72975, R01 AG15819, R01 AG42210), the RUSH Memory and Aging Project (MAP) (R01 AG017917, R01 AG42210Stanford Extreme Phenotypes in AD (R01AG060747), University of Miami Brain Endowment Bank (MBB), University of Miami/Case Western/North Carolina A&T African American (UM/CASE/NCAT) (U01AG052410, R01AG028786), and Wisconsin Registry for Alzheimer’s Prevention (WRAP) (R01AG027161 and R01AG054047).

The four LSACs are: the Human Genome Sequencing Center at the Baylor College of Medicine (U54 HG003273), the Broad Institute Genome Center (U54HG003067), The American Genome Center at the Uniformed Services University of the Health Sciences (U01AG057659), and the Washington University Genome Institute (U54HG003079). Genotyping and sequencing for the ADSP FUS is also conducted at John P. Hussman Institute for Human Genomics (HIHG) Center for Genome Technology (CGT).

Biological samples and associated phenotypic data used in primary data analyses were stored at Study Investigators institutions, and at the National Centralized Repository for Alzheimer’s Disease and Related Dementias (NCRAD, U24AG021886) at Indiana University funded by NIA. Associated Phenotypic Data used in primary and secondary data analyses were provided by Study Investigators, the NIA funded Alzheimer’s Disease Centers (ADCs), and the National Alzheimer’s Coordinating Center (NACC, U24AG072122) and the National Institute on Aging Genetics of Alzheimer’s Disease Data Storage Site (NIAGADS, U24AG041689) at the University of Pennsylvania, funded by NIA. Harmonized phenotypes were provided by the ADSP Phenotype Harmonization Consortium (ADSP-PHC), funded by NIA (U24 AG074855, U01 AG068057 and R01 AG059716) and Ultrascale Machine Learning to Empower Discovery in Alzheimer’s Disease Biobanks (AI4AD, U01 AG068057). This research was supported in part by the Intramural Research Program of the National Institutes of health, National Library of Medicine. Contributors to the Genetic Analysis Data included Study Investigators on projects that were individually funded by NIA, and other NIH institutes, and by private U.S. organizations, or foreign governmental or nongovernmental organizations.

## ROSMAP

We are grateful to the participants in the Religious Order Study, the Memory and Aging Project. This work is supported by the US National Institutes of Health [U01 AG046152, R01 AG043617, R01 AG042210, R01 AG036042, R01 AG036836, R01 AG032990, R01 AG18023, RC2 AG036547, P50 AG016574, U01 ES017155, KL2 RR024151, K25 AG041906-01, R01 AG30146, P30 AG10161, R01 AG17917, R01 AG15819, K08 AG034290, P30 AG10161 and R01 AG11101.

## Author contributions

Study conception and design: M.H.G. and J.E.P-C. Acquisition of study data: W-P.L. and G.D.S. Genotyping of STRs: M.H.G., B.V., and W-P.L. Analyses of STR associations: M.H.G. and J.E.P-C. Writing and figure preparation: led by M.H.G. and J.E.P-C., and reviewed by all authors.

