## Supplementary Figures and Table Legends for "Polygenic burden of short tandem repeat expansions promote risk for Alzheimer’s disease"

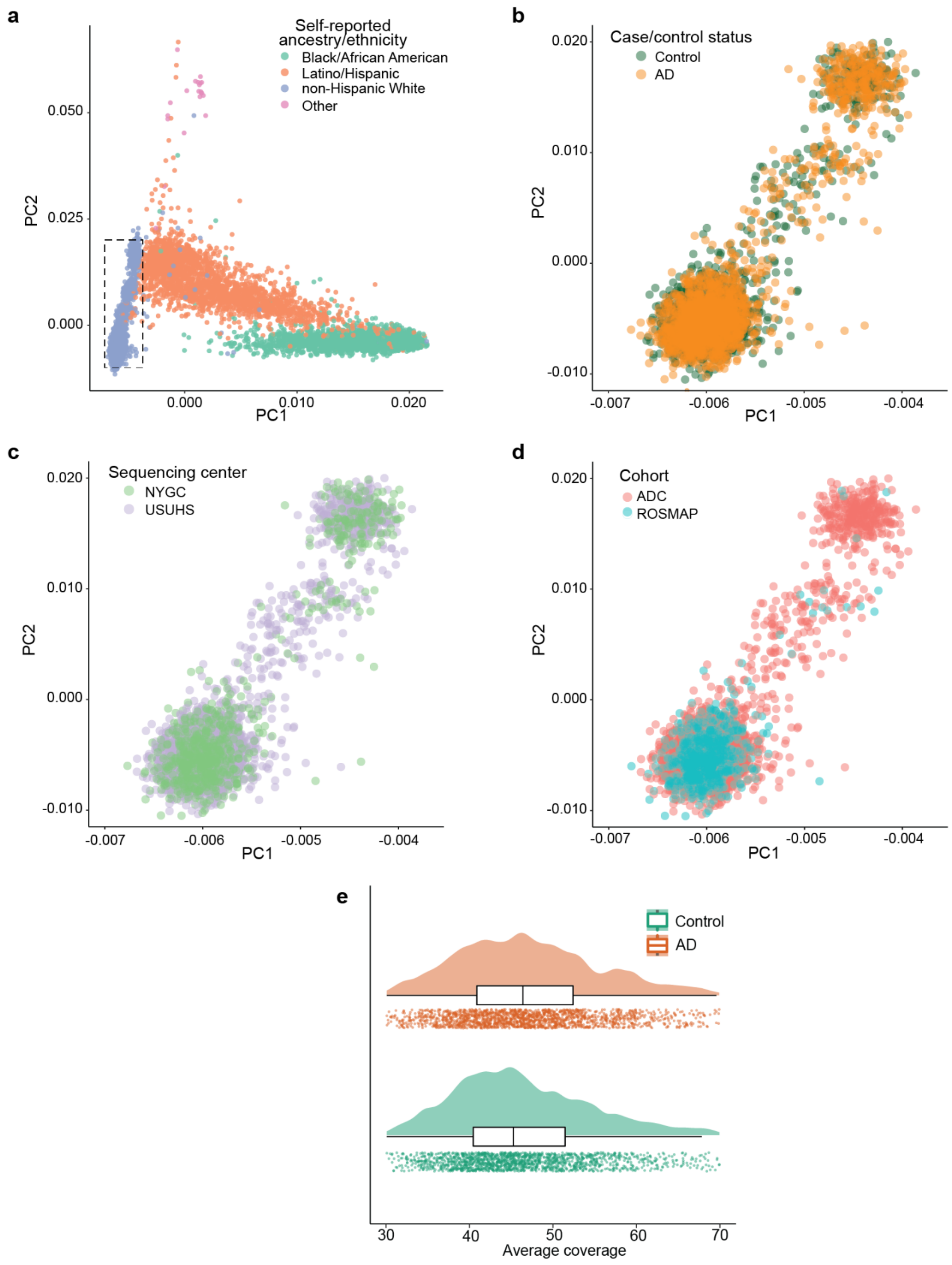


**Supplementary Figure 1: Ancestry and sequencing coverage in whole genome sequencing cohort**. **a,** PCA plot of the n=16,905 individuals in the ADSP, colored by self-identified ethnicity. Dotted box shows the PCA cutoffs used in this study. **b-d,** PCA plot for individuals in this study colored by case/control status **b,** cohort **c,** or sequencing center **d**. **e,** Raincloud plot showing distribution of average sequencing coverage for each individual in this study. Each individual is represented by one point.

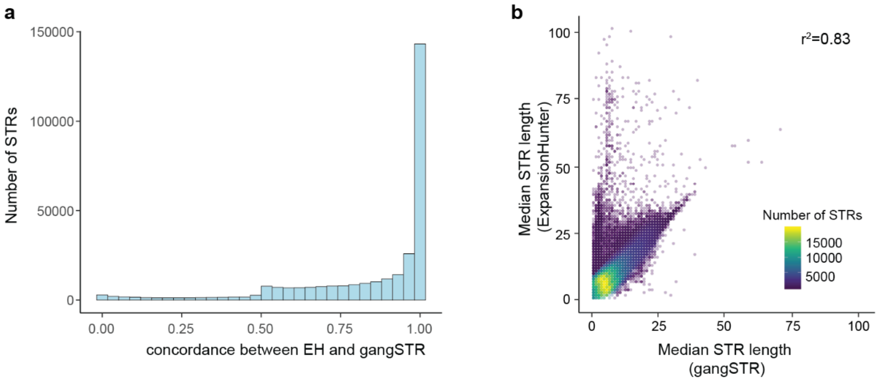


**Supplementary Figure 2: Comparison of ExpansionHunter and gangSTR algorithms for genotyping STRs. a,** Histogram of concordance between ExpansionHunter and gangSTR genotypes across STRs. Height of each bar is the number of STRs falling within a given concordance bin. **b,** Correlation between gangSTR genotypes and ExpansionHunter genotypes. Each point represents the median tract length for an STR as measured by gangSTR (x-axis) and ExpansionHunter (y-axis).

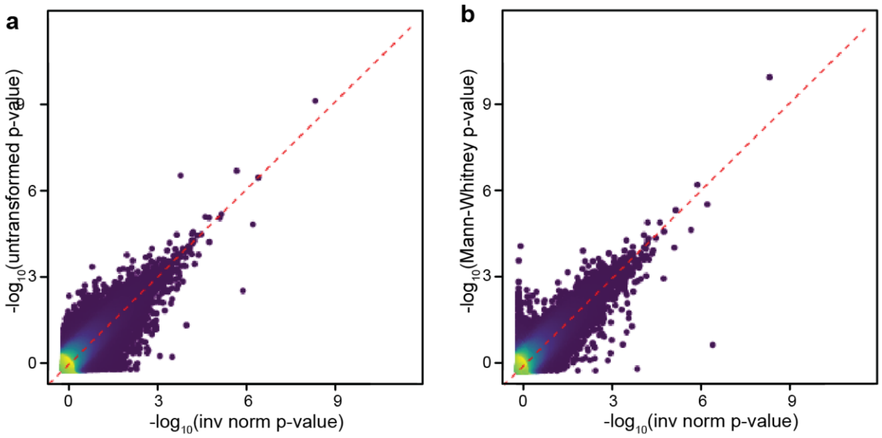


**Supplementary Figure 3: Comparison of statistical models for single STR associations with AD risk. a,** Scatterplot for -$\log_{10}$(p-values) for a logistic model without inverse normal transformation (y-axis) versus with inverse normal transformation (x-axis). **b,** Scatterplot for -$\log_{10}$(p-values) for a non-parametric model (Mann-Whitney test) without covariates (y-axis) versus with inverse normal transformation (x-axis).

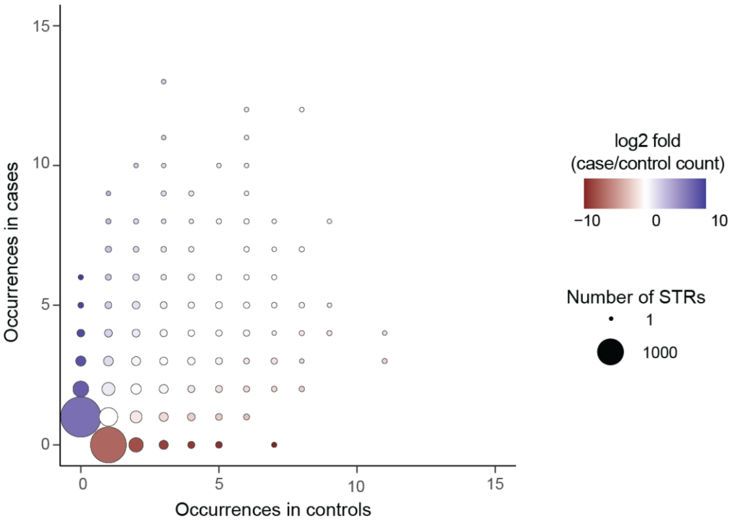


**Supplementary Figure 4: Recurrence of STR expansions in AD cases and controls.** Plot shows the number of times each STR expansion was observed in cases versus controls. Size of each circle reflects the number of STRs observed in a given number of case and control individuals. Colors of circles reflect the $\log_2$fold of the ratio of times an STR was observed in cases versus controls.

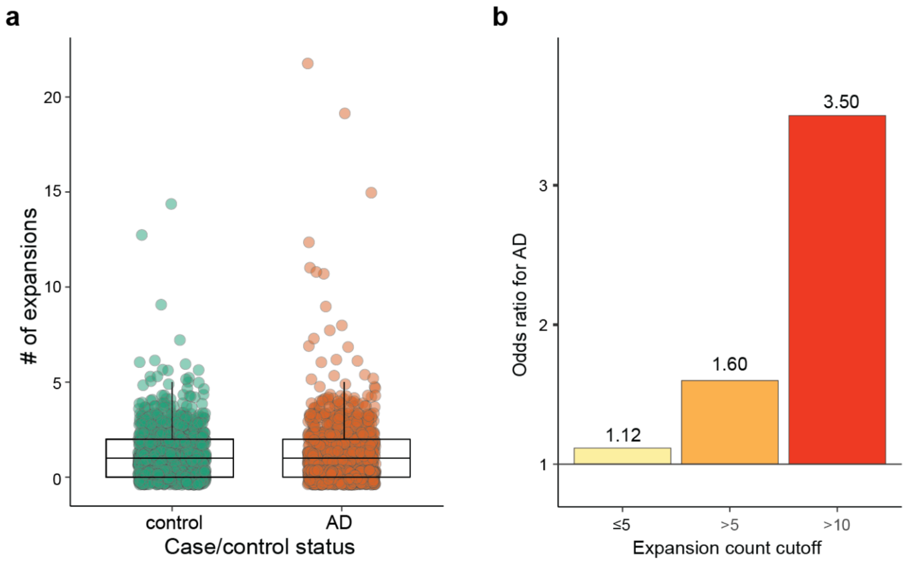


**Supplementary Figure 5: Burden of STR expansions based on genotyping using gangSTR. a,** Number of STR expansions per individual identified by DBSCAN using gangSTR genotypes in controls (green) versus cases (orange). Each point represents one individual. **b,** Odds ratio for AD case/control status for individuals carrying varying numbers of STR expansions as identified using gangSTR. Odds ratios > 1.0 represent higher odds of having AD.

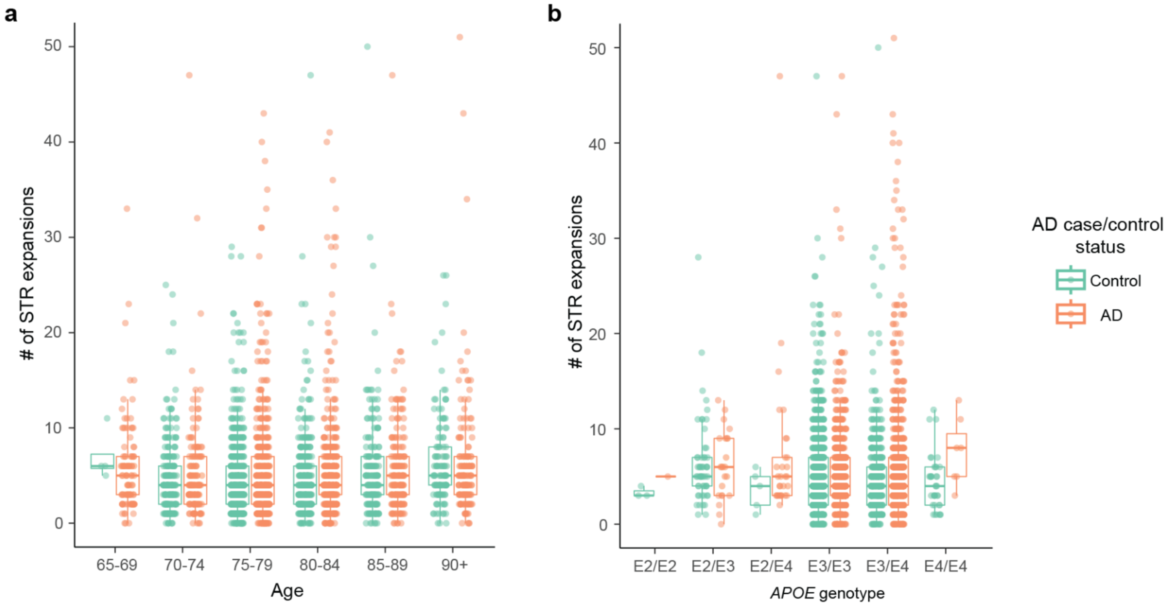


**Supplementary Figure 6: Association of age and *APOE* genotype with STR expansion burden. a,** Number of STR expansions for controls (green) or cases (orange) for individuals of different diploid *APOE* genotypes. **b,** Number of STR expansions for controls (green) or cases (orange) for individuals at different ages. Each point represents one individual.

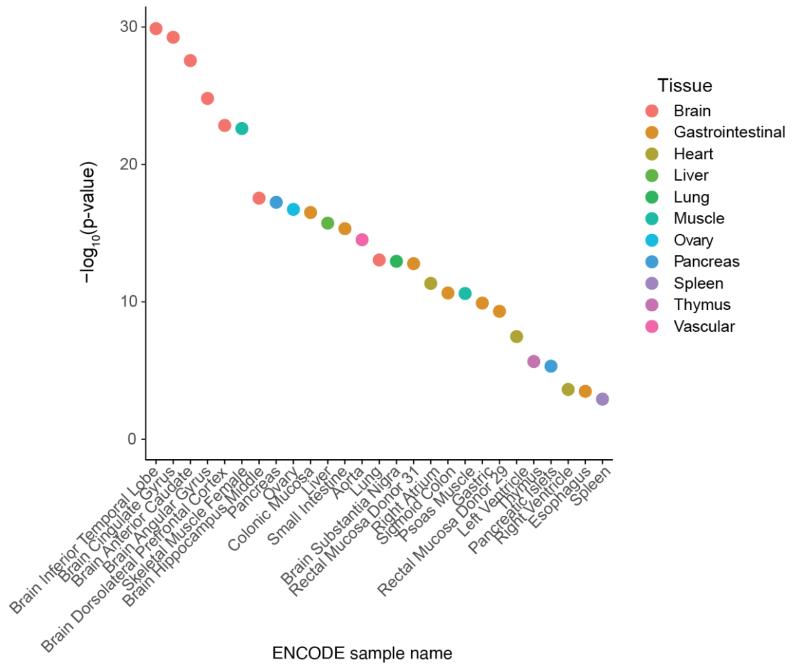


**Supplementary Figure 7: Enrichment of AD STR expansions in "flanking TSS" regions**. Enrichments of AD STR expansions in chromatin state 2 ("flanking TSS") across tissues in ENCODE. Height of the points reflect the -log10 of the enrichment p-value. Points are colored by organ.

**Supplementary Table Legends**

**Supplementary Table 1: Catalog of polymorphic STRs.** Genomic coordinates, number of repeats in reference genome (reference tract length), and motif are shown based on GRCh38 reference genome. Segmental duplication column indicates whether the STR is within a segmentally duplicated region (1 if within a segmental duplication, 0 if not). Genomic annotations are listed for each STR, as well as the distance to nearest gene and the corresponding gene Ensembl ID and gene symbol.

**Supplementary Table 2: Results of single STR association analysis with AD risk.** Genomic coordinates, number of repeats in reference genome (reference tract length), and motif are shown based on GRCh38 reference genome. Effect shows the $\beta$ from the linear regression, with positive values reflecting longer tract lengths in AD cases and negative values reflecting longer tract lengths in controls. Association pval is the linear regression p-value. Alleles AD and alleles control show the STR tract lengths and the number of individuals with that tract length among cases and controls, respectively.

**Supplementary Table 3: Results of hypergeometric test for STR expansions in AD.** Genomic coordinates, number of repeats in reference genome (reference tract length), and motif are shown based on GRCh38 reference genome. The total number of cases and total number of controls that could be genotyped for that STR are listed. The number of cases with an expansion of the STR (case_count_min_XXXrep), number of controls with an expansion of the STR (control_count_min_XXXrep) and the p-value of the hypergeometric test (fisher_pval_min_XXXrep) are shown, where XXX is the STR tract length threshold (either ≥1, ≥5, ≥10, or ≥20 repeat units longer than the GRCh38 reference genome).

**Supplementary Table 4: List of STR expansions identified by DBSCAN.** Genomic coordinates, number of repeats in reference genome (reference tract length), and motif are shown based on GRCh38 reference genome for each STR expansion identified by DBSCAN. The tract length of the expansion and whether it was seen in a case or control individual is also listed.

**Supplementary Table 5: Number of expansions identified by DBSCAN per individual.** Each row represents a separate individual. The case/control status, age, Braak stage, sequencing coverage, and total number of expansions identified by DBSCAN based on ExpansionHunter genotypes in that individual are shown.
